# Deaths in Children and Young People in England following SARS-CoV-2 infection during the first pandemic year: a national study using linked mandatory child death reporting data

**DOI:** 10.1101/2021.07.07.21259779

**Authors:** C Smith, D Odd, R Harwood, J Ward, M Linney, M Clark, D Hargreaves, SN Ladhani, E Draper, PJ Davis, SE Kenny, E Whittaker, K Luyt, RM Viner, LK Fraser

## Abstract

**Background:** Deaths in children and young people (CYP) following SARS-CoV-2 infection are rare. Quantifying the risk of mortality is challenging because of high relative prevalence of asymptomatic and non-specific disease manifestations. Therefore, it is important to differentiate between CYP who have died of SARS-CoV-2 and those who have died of an alternative disease process but coincidentally tested positive.

**Methods:** During the pandemic, the mandatory National Child Mortality Database (NCMD) was linked to Public Health England (PHE) testing data to identify CYP (<18 years) who died with a positive SARS-CoV-2 test. A clinical review of all deaths from March 2020 to February 2021 was undertaken to differentiate between those who died of SARS-CoV-2 infection and those who died of an alternative cause but coincidentally tested positive. Then, using linkage to national hospital admission data, demographic and comorbidity details of CYP who died of SARS-CoV-2 were compared to all other deaths. Absolute risk of death was estimated where denominator data were available.

**Findings:** 3105 CYP died from all causes during the first pandemic year in England. 61 of these deaths occurred in CYP who tested positive for SARS-CoV-2. 25 CYP died of SARS-CoV-2 infection; 22 from acute infection and three from PIMS-TS. 99·995% of CYP with a positive SARS-CoV-2 test survived. The 25 CYP who died of SARS-CoV-2 equates to a mortality rate of 2/million for the 12,023,568 CYP living in England. CYP >10 years, of Asian and Black ethnic backgrounds, and with comorbidities were over-represented compared to other children.

**Interpretation:** SARS-CoV-2 is very rarely fatal in CYP, even among those with underlying comorbidities. These findings are important to guide families, clinicians and policy makers about future shielding and vaccination.

**Funding:** RH is in receipt of a fellowship from Kidney Research UK. JW is in receipt of a Medical Research Council Fellowship. LF is in receipt of funding from Martin House Childrens Hospice.

## Introduction

Identifying Children and Young People (CYP) at risk of severe illness and death following SARS-CoV-2 infection is essential to guide families, clinicians and policy makers about future shielding policies, school attendance and vaccine prioritisation.

SARS-CoV-2 infection is usually mild and asymptomatic in CYP.^1,2^ Therefore, CYP have comprised a very low proportion of all hospitalisations and deaths from COVID-19 globally.^3^ The clinical manifestations of COVID-19 in CYP are different to those amongst adults.^1^ While many CYP present with the typical fever, cough and shortness of breath, they also present with broader non-specific symptoms including abdominal pain, nausea, headache and sore throat.^1,3^ This, in combination with a mild or asymptomatic phenotype^2^, provides a challenge for describing how SARS-CoV-2 directly affects CYP.

Severe illness and death in CYP is rare and can be due to either acute COVID-19 or Paediatric Inflammatory Multisystem Syndrome Temporally Associated with SARS-CoV-2 (PIMS-TS).^2,4^ PIMS-TS, also called Multi-System Inflammatory Syndrome in Children (MIS-C), is a rare syndrome characterised by persistent fever, inflammation (neutrophilia, lymphopaenia, and raised CRP) and evidence of single or multi-organ dysfunction.^5^ As death from acute COVID-19 or PIMS-TS amongst CYP is extremely rare^3,6,7^ those that have died have been poorly characterised. Further, it remains unclear to what extent these rare deaths relate directly to the pathological processes of COVID-19 or whether CYP who died from alternative causes were coincidentally SARS-CoV-2 positive around the time of death. This issue is made more difficult by the very high prevalence of asymptomatic infection at times of high prevalence, with reported prevalence rates up to 4-6% of UK CYP during December 2020.^8^ The distinction between those who died of SARS-CoV-2 infection and those who died of an alternative cause with a coincidental positive SARS-CoV-2 test, is important for understanding which CYP are truly at higher risk for severe disease or death.

To answer this important question required detailed examination of all deaths in a large population, going beyond simple cause of death registration, to review the contribution of SARS-CoV-2 to death. We used detailed clinical data in the National Child Mortality Database (NCMD)^9^, a comprehensive and unique mandatory national dataset of deaths <18 years of age, to review the contribution of SARS-CoV-2 to death. We compared the characteristics of those who died of SARS-CoV-2 to those who died from all other causes during the first pandemic year.

If higher risk groups are identified, they may benefit from vaccination and/or protective ‘shielding’ at times of high prevalence, whereas ‘shielding’ based upon erroneous assumptions of vulnerability is likely to cause significant secondary harms. Similarly, risks from the disease need to be weighed against risks of vaccination in informing vaccination policy.

## Method

### Population

The cohort investigated in this study is all CYP <18 years of age, who died in England between 1^st^ March 2020 and the 28^th^ February 2021.^9^ The aim of this study was to identify CYP in which SARS-CoV-2 contributed to death, i.e. they died of SARS-CoV-2 infection.

### Data Collection

The NCMD is a mandatory system that records all deaths in CYP <18 years of age in England, since it began in April 2019,^9^ and includes demographic and clinical data of the events leading up to death.

In this analysis, demographic details included age (coded as 0-27 days, 28-364 days, 1-4 years, 5-9 years, 10-14 years and 15-17 years), sex, ethnicity (coded as Asian or Asian British, Black or African or Caribbean or Black British, Mixed, Multiple or Other, or White^10^) and deprivation (see supplementary information).^11, 12^

### Data linkage

To ensure comprehensive identification of comorbidities, NCMD data were linked to the preceding five years (March 2015 onwards) of national admitted patient care Secondary Uses Service (SUS) data for England^13^ and to the national Paediatric Intensive Care Audit Network (PICANet) data. A validated list of ICD-10 codes was used to identify CYP with chronic co-morbidities^14^ and life-limiting conditions^15^. Of note, the chronic disease list for cardiac conditions was modified to remove ‘I46-Cardiac Arrest’ and ‘I51-Complications and ill-defined descriptions of heart disease’ as these are acute presentations of cardiac disease and likely to represent PIMS-TS rather than pre-existing comorbidity. We also identified CYP with chronic comorbidities in two body systems and with the following single diagnoses: asthma, diabetes, epilepsy, sickle cell disease and trisomy 21.

### SARS-CoV-2 Data

During the pandemic, the NCMD was linked by NHS number to Public Health England (PHE) Pillar 1 and Pillar 2 testing data^16^ to identify all CYP who died with a positive SARS-CoV-2 test. Pillar 1 testing occurs in health and care settings, while Pillar 2 testing occurs in the community^1616^, both started in March 2020. The NCMD contributed to modification of the protocol for sudden unexpected deaths in CYP to include post-mortem testing for SARS-CoV-2^17^. All CYP who died with a positive SARS-CoV-2 test were included, regardless of the time interval between positive test and death. This is different to the definition used for reporting adult deaths to ensure all potential cases were identified for review and to optimise capture of possible PIMS-TS cases. In addition, the NCMD coding team identified potential cases of PIMS-TS (see supplementary material).

### Identifying CYP who died of SARS-CoV-2

Clinical records of all CYP who died with a positive SARS-CoV-2 test were reviewed to identify if SARS-CoV-2 clearly, probably, possibly or unlikely contributed to death (Figure 1 and supplementary material).

**Figure 1.**
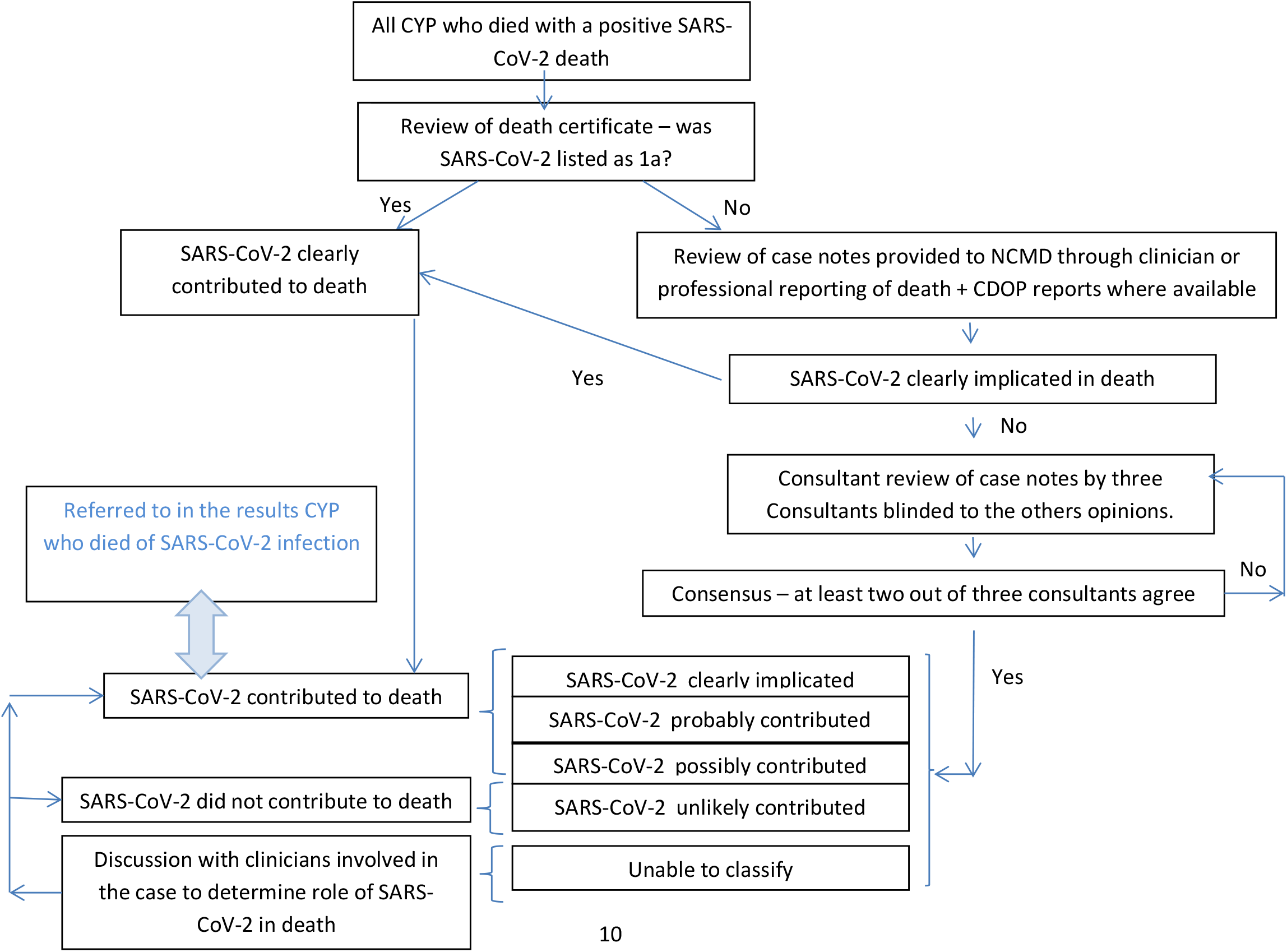
Approach to determine if SARS-CoV-2 contributed to death or if it was a co-incidental finding

### Statistical Analysis

The group of CYP who died of SARS-CoV-2 were compared to CYP who died from all other causes using summary statistics and differences between groups were compared using Chi2 or Fishers’ Exact test if small numbers. The comparator cohort, death from all other causes, included CYP who tested positive for SARS-CoV-2, but died of another cause.

The absolute risk of death was calculated for the whole population and for demographic groups in which denominator data were available. The quality of available data on the number of CYP in the population with comorbidities was variable. We have used estimates for comorbidity groups, where we have enough confidence in the data, to derive estimated absolute risk. This data came from a range of sources and is referenced in Table 3.

Infection fatality rate was calculated using the number of CYP infected with SARS-CoV-2 during the same time period (March 2020 to February 2021) estimated through PHE modelling data.^18^ This was chosen rather than the absolute number of positive SARS-CoV-2 tests as CYP may test positive more than once, and many CYP were not tested in the first wave of the pandemic. Mortality rate was calculated using a population of 12,023,568 CYP living in England^19^ during the study year.

### Small number reporting, information governance and legal basis

See supplementary information.^20,21^

## Results

Between March 2020 and February 2021, 3105 CYP in England died of all causes. Of these, 61 CYP had a positive SARS-CoV-2 test (5 deaths every 30 days) and 3044 died from all other causes (250 deaths every 30 days).

Clinical records of the 61 CYP who died with a positive SARS-CoV-2 test were reviewed to identify if SARS-CoV-2 contributed to death. 25 (41%) of the 61 CYP died of SARS-CoV-2 (2 deaths every 30 days), including 22 with acute COVID-19 and three with PIMS-TS. The other 36 (59%) of the 61 CYP were categorised as SARS-CoV-2 did not contribute to death (Table 1, Figure 1, Figure 2).

**Table 1.** Demographic details for Children and Young People (CYP) who died between March 2020 to February 2021 from all causes, and the 61 CYP who died with a positive SARS-CoV-2 test, split by the likely cause of death.

| All Deaths |  | All deaths March 2020 - February 2021 (n=3105) |  |  |  |  |  |  |  | Comparison of frequencies* |
| --- | --- | --- | --- | --- | --- | --- | --- | --- | --- | --- |
|  |  | Reference Population (n=3080) - All other causes of death |  |  |  |  | Died of COVID-19/PIMS-TS (n=25) |  |  |  |
|  |  | Died without positive test for SARS-CoV-2 (N=3044) |  | Incidental positive SARS-CoV-2 test at death (n=36) |  |  |  |  |  |  |
|  |  | Number | Percentage | Number | Percentage | Est Rate/100,000 person years (95%Poisson CI) | Number | Percentage | Est Rate/100,000 person years (95%Poisson CI) | P |
|  |  | 3044 | 100% | 36 | 100% | 25.5 (24.7-26.5) | 25 | 100% | 0.2 (0.1-0.3) |  |
| Age | 0 - 27 days | 1388 | 45.6% | 3 | 8.3% | 289.3 (276.8-302.1) | 0 | 0.0% | 0.3 (0.0-11.0) | <0.001 |
|  | 28 - 364 days | 616 | 20.2% | 11 | 30.60% |  | 2 | 8.0% |  |  |
|  | 1 - 4 years | 281 | 9.2% | 4 | 11.1% | 10.4 (9.2-11.6) | 0 | 0.0% | 0.00 (-) |  |
|  | 5 - 9 years | 191 | 6.3% | 4 | 11.1% | 5.7 (4.9-6.6) | 5 | 20.0% | 0.1 (0.0-0.3) |  |
|  | 10 - 14 years | 252 | 8.3% | 6 | 16.7% | 7.6 (6.7-8.6) | 9 | 36.0% | 0.3 (0.1-0.5) |  |
|  | 15 - 17 years | 316 | 10.4% | 8 | 22.2% | 17.3 (15.5-19.3) | 9 | 36.0% | 0.5 (0.2-0.9) |  |
| Sex | Female | 1667 | 56.0% | 18 | 50.0% | 22.4 (21.2-23.7) | 12 | 48.0% | 0.2 (0.1-0.4) | 0.28 |
|  | Male | 1308 | 44.0% | 18 | 50.0% | 27.1 (25.9-28.5) | 13 | 52.0% | 0.2 (0.1-0.3) |  |
| Ethnicity | Asian or Asian British | 471 | 17.3% | 10 | 28.6% | 31.3 (28.6-34.2) | 9 | 36.0% | 0.6 (0.3-1.1) | <0.001 |
|  | Black or Black British | 241 | 8.9% | 4 | 11.4% | 37.8 (33.2-42.9) | 5 | 20.0% | 0.8 (0.3-1.8) |  |
|  | Mixed | 169 | 6.2% | 1 | 2.9% | 23.3 (19.9-27.1) | 4 | 16.0% | 0.5 (0.1-1.4) |  |
|  | Other | 74 | 2.7% | 0 | 0% | 49.8 (39.1-62.5) | 0 | 0.0% | 0.00 (-) |  |
|  | White | 1764 | 64.9% | 20 | 57.1% | 19.7 (18.8-20.6) | 7 | 28.0% | 0.1 (0.0-0.2) |  |
| Deprivation Category | 1 | 985 | 32.6% | 15 | 41.7% | 34.8 (32.7-37.0) | 9 | 36.0% | 0.3 (0.1-0.6) | 0.99 |
|  | 2 | 687 | 22.7% | 8 | 22.2% | 27.9 (25.9-30.1) | 6 | 24.0% | 0.2 (0.1-0.5) |  |
|  | 3 | 615 | 20.3% | 6 | 16.7% | 27.4 (25.3-29.6) | 5 | 20.0% | 0.2 (0.1-0.5) |  |
|  | 4 | 402 | 13.3% | 3 | 8.3% | 18.7 (16.9-20.6) | 3 | 12.0% | 0.1 (0.0-0.4) |  |
|  | 5 | 337 | 11.1% | 4 | 11.1% | 15.3 (13.8-17.1) | 2 | 8.0% | 0.1 (0.0-0.3) |  |

Values are n(%) or median (IQR) as appropriate
\* Comparison of frequencies seen between CYP likely dying of SAR-CoV-2 and all other infants

**Figure 2:**
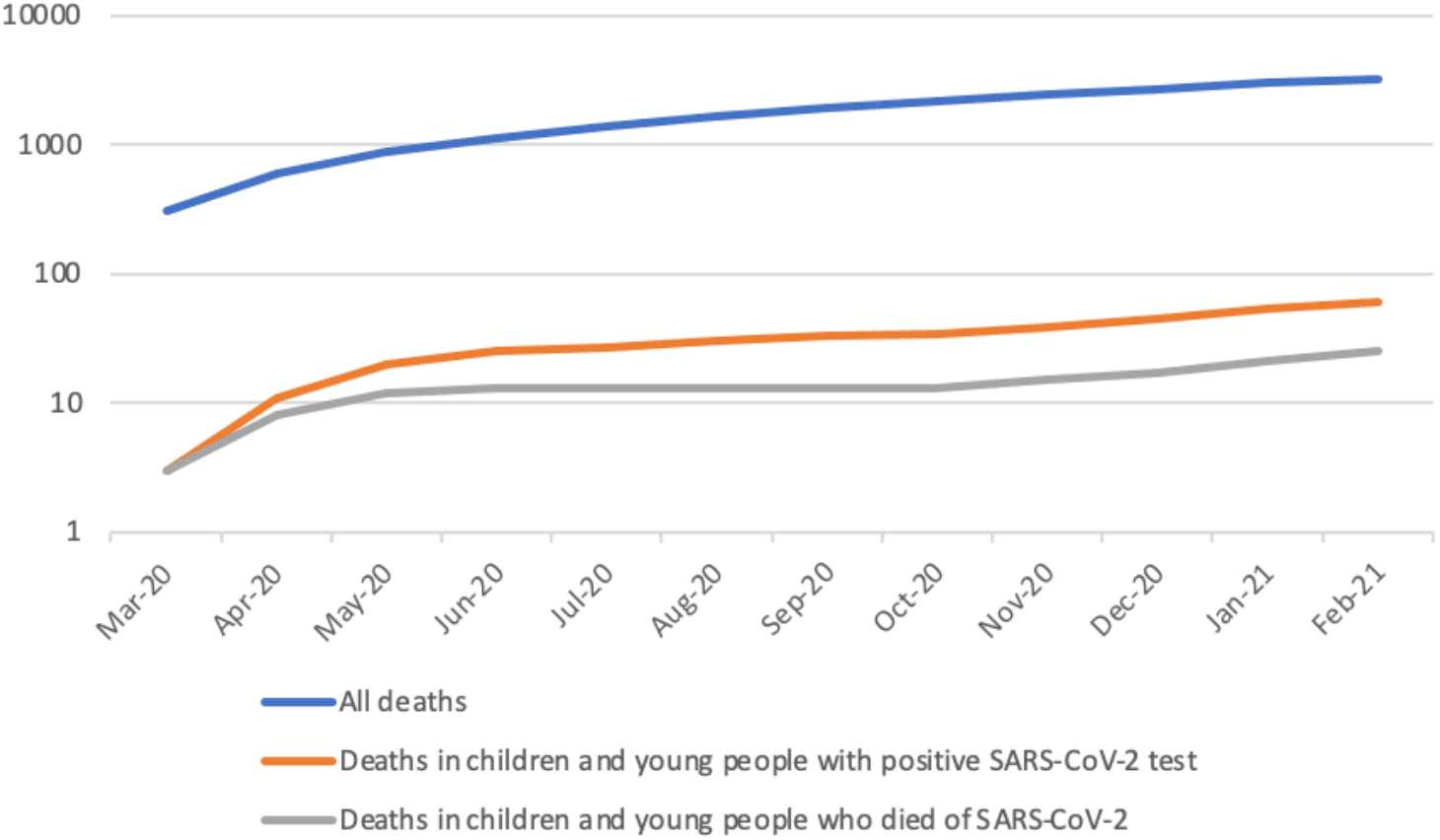
Cumulative number of deaths for all CYP who died with a positive SARS-CoV-2 test, who died of SARS-CoV-2 and who died of all other causes

There were an estimated 469,982 CYP infected with SARS-CoV-2 in England from March 2020 to February 2021, giving an infection fatality rate of 5 per 100,000 CYP (0·005%) and, based on a population of 12,023,568, a mortality rate of 2 per million CYP (0·0002%).

### Demographics (Table 1, Figures 3 and 4)

CYP who died of SARS-CoV-2 (n=25) were older than those who died from all other causes (n=3080) in the same time period (Figure 3). 18/25 (72%) young people who died of SARS-CoV-2 were aged 10 years or over, compared to 19% in the deaths from all other causes (chi2 59·7, p <0·001). All three deaths in CYP who died of PIMS-TS were aged 10-14 years. The sex distribution was equally split between males and females (12 (48%) and 13 (52%) respectively) and did not differ from the deaths from all other causes (chi2 0·64, p=0·28). A greater proportion of CYP from an Asian (36% cf 16%) and Black (20% cf 8%) ethnicity died of SARS-CoV-2 compared to deaths from all other causes (chi2 17·9, p<0·001). There was no significant difference in the deprivation categories between CYP who died of SARS-CoV-2 and deaths from all other causes (chi2 0·35, p=0·99) although more CYP from more deprived areas died in both groups.

**Figure 3:**
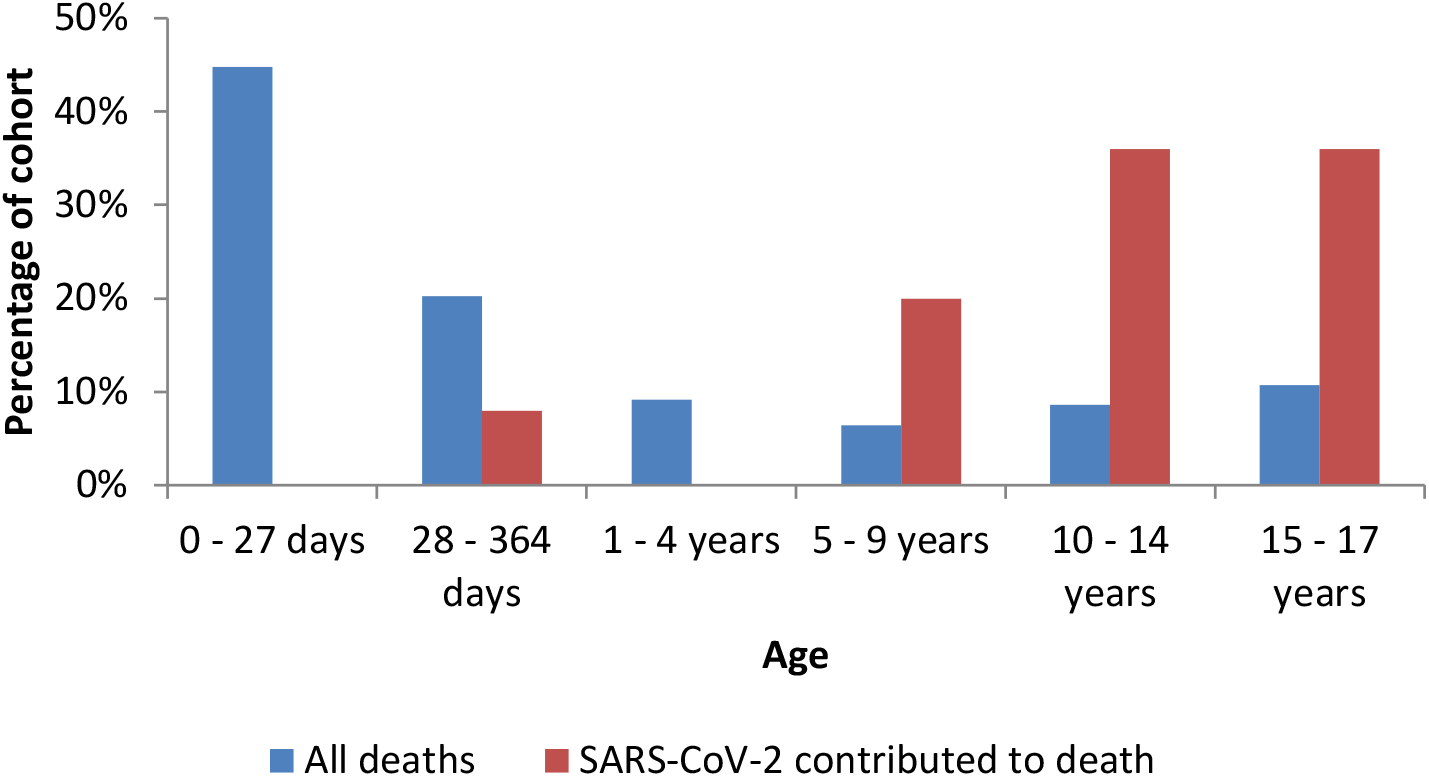
Age group of CYP for all deaths (n=3080) and CYP who died of SARS-CoV-2 (n=25)

**Figure 4:**
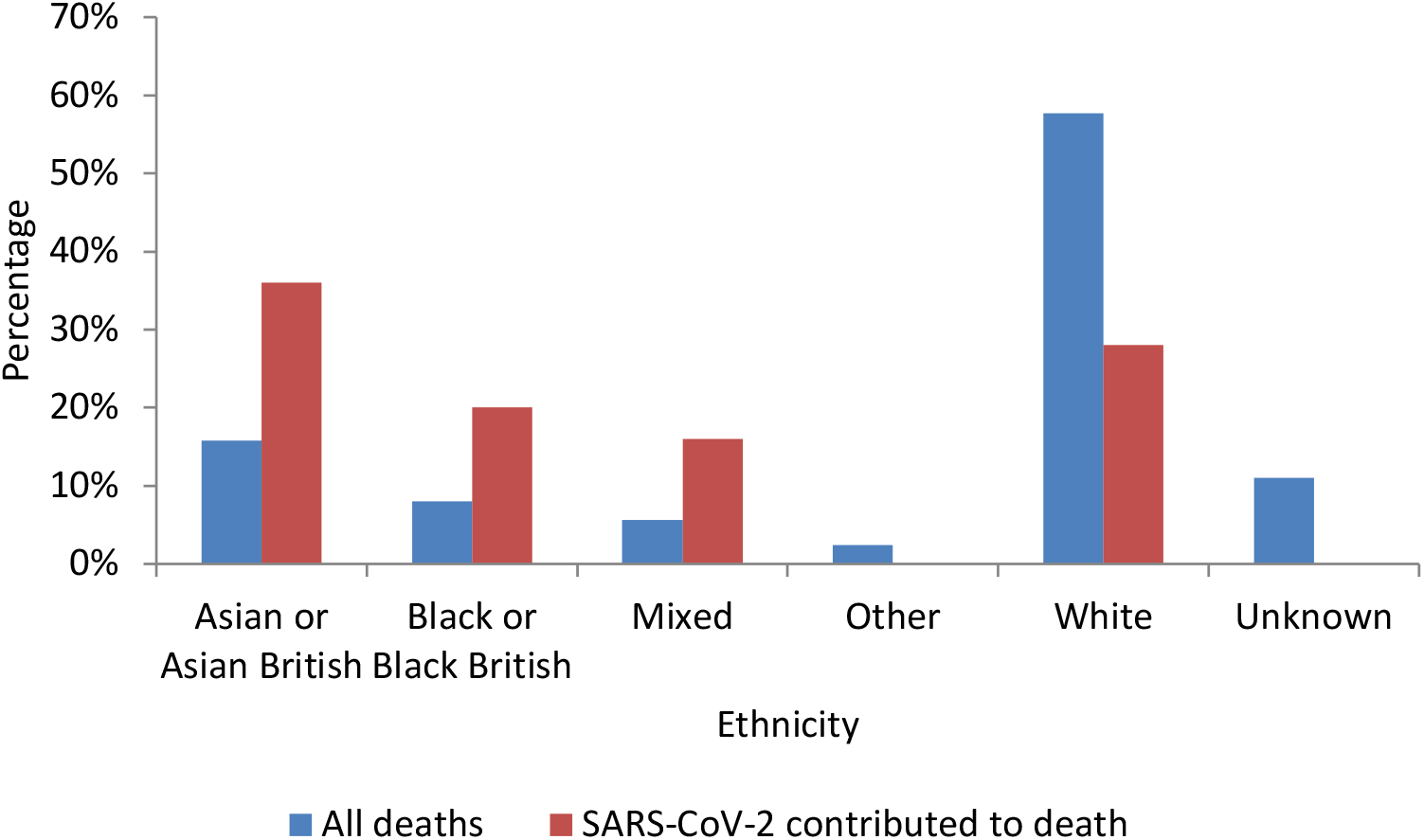
Ethnic group of CYP for all deaths (n=3080) and CYP who died of SARS-CoV-2 (n=25)

The mortality rate in CYP who died of SARS-CoV-2 was 0·2 per 100,000 (95%CI 0·1-0·3) compared to 25·5 per 100,000 (95%CI 24·7-26·5) for all other causes of death. Although the proportion of CYP from Asian and Black ethnic groups who died of SARS-CoV-2 was higher, their absolute risk of death from SARS-CoV-2 was still extremely low at 0·6 per 100,000 (95%CI 0·3-1·1) and 0·8 per 100,000 (95%CI 0·3-1·8) respectively. Similarly the proportion of CYP aged 10-14 years and 15-17 years who died of SARS-CoV-2 was higher, however their absolute risk of death from SARS-CoV-2 was still extremely low at 0·3 per 100,000 (95%CI 0·1-0·5) and 0·5 (95%CI 0·2-0·9) per 100,000 respectively.

### Co-morbidities (Table 2, Table 3)

A similar proportion of the 25 CYP who died of SARS-CoV-2 (n=19, 76%) and the 3080 deaths from all other causes (n=2267, 74%) (chi2 0·004, p=0·60) had a chronic underlying health condition. Significantly more CYP who died of SARS-CoV-2 had a life-limiting condition (n=15, 60%) compared to deaths from all other causes (n=988, 32%) (chi2 8·5, p=0·005). 64% (n=16) of the 25 CYP who died of SARS-CoV-2 had comorbidities in two or more body systems compared to 45% (n=1373) of the CYP who died from all other causes (chi2 5·5, p=0·14).

**Table 1.** Co-morbidity details for CYP who died between March 2020 to February 2021 from all causes, and the 61 CYP who died with a positive SARS-CoV-2 test, split by the likely cause of death.

| All deaths |  | All deaths March 2020 - February 2021 (n=3105) |  |  |  |  |  | Comparison of frequencies* |
| --- | --- | --- | --- | --- | --- | --- | --- | --- |
|  |  | Reference Population (n=3080) |  |  |  | Died of COVID-19/PIMS-TS (n=25) |  |  |
|  |  | All causes (n=3044) |  | Incidental positive SARS-CoV-2 test at death(n=36) |  |  |  |  |
|  |  | Number | Percentage | Number | Percentage | Number | Percentage |  |
| Life-limiting condition | Yes | 974 | 32.0% | 14 | 38.9% | 15 | 60.0% | 0.005 |
|  | No | 2027 | 66.6% | 22 | 61.1% | 10 | 40.0% |  |
| Chronic condition | Yes | 2238 | 73.5% | 29 | 80.6% | 19 | 76.0% | 0.6 |
|  | No | 716 | 23.5% | 7 | 19.4% | 6 | 24.0% |  |
| Chronic Condition Details | Cardiology (Non-congenital) | 458 | 15.0% | 8 | 22.2% | 8 | 32.0% | 0.02 |
|  | Cardiology (Congenital) | 667 | 21.9% | 10 | 27.8% | 1 | 4.0% | 0.03 |
|  | Dermatology | 14 | 0.5% | 0 | 0.0% | 0 | 0.0% | - |
|  | Endocrine | 29 | 1.0% | 1 | 2.8% | 1 | 4.0% | 0.22 |
|  | ENT | 70 | 2.3% | 5 | 13.9% | 10 | 40.0% | <0.001 |
|  | Gastroenterology | 467 | 15.3% | 18 | 50.0% | 5 | 20.0% | 0.56 |
|  | Genetic | 88 | 2.9% | 1 | 2.8% | 8 | 32.0% | <0.001 |
|  | Haematological | 287 | 9.4% | 12 | 33.3% | 2 | 8.0% | 0.81 |
|  | Immunological | 19 | 0.6% | 2 | 5.6% | 1 | 4.0% | 0.16 |
|  | Infectious disease | 15 | 0.5% | 0 | 0.0% | 0 | 0.0% | - |
|  | Metabolic | 181 | 5.9% | 7 | 19.4% | 4 | 16.0% | 0.07 |
|  | Musculoskeletal | 142 | 4.7% | 5 | 13.9% | 4 | 16.0% | 0.03 |
|  | Neurological | 1194 | 39.2% | 24 | 66.7% | 13 | 52.0% | 0.29 |
|  | Oncology | 190 | 6.2% | 8 | 22.2% | 1 | 4.0% | 0.51 |
|  | Renal | 300 | 9.9% | 6 | 16.7% | 2 | 8.0% | 0.74 |
|  | Reproductive system | 9 | 0.3% | 1 | 2.8% | 0 | 0.0% | - |
|  | Respiratory | 474 | 15.6% | 10 | 27.8% | 12 | 48.0% | <0.001 |
|  | Rheumatology | 4 | 0.1% | 1 | 2.8% | 0 | 0.0% | - |
|  | Trauma | 12 | 0.4% | 2 | 5.6% | 0 | 0.0% | - |

| All deaths |  | Reference Population (n=3080) |  |  |  | Died of COVID-19/PIMS-TS (n=25) |  | Comparison of frequencies* |
| --- | --- | --- | --- | --- | --- | --- | --- | --- |
|  |  | All causes (n=3044) |  | Incidental positive SARS-CoV-2 test at death(n=36) |  |  |  |  |
|  |  | Number | Percentage | Number | Percentage | Number | Percentage | p |
| Number of comorbidities | 0 | 716 | 23.5% | 13 | 21.3% | 6 | 24.0% | 0.14 |
|  | 1 | 906 | 29.8% | 7 | 11.5% | 3 | 12.0% |  |
|  | 2 or more | 1332 | 43.8% | 41 | 67.2% | 16 | 64.0% |  |
|  | Unknown | 90 | 3.0% | 0 | 0.0% | 0 | 0.0% |  |
|  | Total | 3044 | 100.0% | 61 | 100.0% | 25 | 100.0% | - |
| Comorbidity combinations | Neurological & Respiratory | 318 | 10.4% | 17 | 27.9% | 8 | 32.0% | <0.001 |
|  | Neurological & Cardiology | 559 | 18.4% | 15 | 24.6% | 3 | 12.0% | 0.61 |
|  | Respiratory & Cardiology | 270 | 8.9% | 12 | 19.7% | 3 | 12.0% | 0.49 |
| Single diagnoses | Asthma** | 58 | 1.9% | 5 | 8.2% | 3 | 12.0% | 0.02 |
|  | Type 1 Diabetes | 9 | 0.3% | 0 | 0.0% | 0 | 0.0% | - |
|  | Epilepsy** | 199 | 6.5% | 7 | 11.5% | 7 | 28.0% | <0.001 |
|  | Sickle cell disease | 1 | 0.0% | 1 | 1.6% | 1 | 4.0% | 0.02 |
|  | Trisomy 21 | 38 | 1.2% | 0 | 0.0% | 0 | 0.0% | - |

Values are n(%)
\* Comparison of frequencies seen between CYP likely dying of SARS-CoV-2 and all other CYP using chi2 or fishers exact if cell <5
\*\*There were zero deaths in CYP with an isolated diagnosis of asthma or epilepsy. All the deaths in CYP with asthma or epilepsy had an additional neurological comorbidity.

**Table 3:** Estimated Mortality Rates by selected diagnostic groups

|  | Estimated population at risk | Reference Population, all other causes (n=3080) |  | Died of COVID-19/PIMS-TS (n=25) |  |
| --- | --- | --- | --- | --- | --- |
|  |  | Number | Est Rate/100,000 person years<br>(95%Poisson CI) | Number | Est Rate/100,000 person years<br>(95%Poisson CI) |
| **All children** | 12,118,268<sup>19</sup> | 3080 | 25.4 (24.5,26.3) | 25 | 0.2 (0.1,0.3) |
| **Oncology** | 1,065<sup>27</sup> | 137 | 12864 (10800,15207) | 1 | 93.9 (2.4,523.2) |
| **Life-limiting Neurodisability** | 14,626<sup>15</sup> | 357 | 2441 (2194,2707) | 13 | 88.9 (47.3,152.0) |
| **Life-limiting condition** | 86,625<sup>15</sup> | 974 | 1124 (1054,1197) | 15 | 11.5 (5.6,21.2) |
| **Cardiology (Congenital)** | 90,000<sup>28</sup> | 458 | 508.9 (463.3,557.7) | 1 | 8.9 (3.8,17.5) |
| **Epilepsy\*\*** | 90,000<sup>29</sup> | 199 | 214 (185,246) | 7 | 7.5 (3.0,15.5) |
| **Asthma\*\*** | 1,100,000<sup>30</sup> | 58 | 5.3 (4.0,6.8) | 3 | 0.3 (0.06,0.8) |

\*\*There were zero deaths in CYP with an isolated diagnosis of asthma or epilepsy. All the deaths in CYP with asthma or epilepsy had an additional neurological comorbidity

Six (24%) of the 25 CYP who died of SARS-CoV-2 appeared to have no underlying health conditions similar to 24% (729 of the 3080 CYP) who died of all other causes.

Neurological conditions were the commonest comorbidity in both the CYP who died of SARS-CoV-2 (n=13/25, 52%) and the CYP who died of all other causes (n=1218/3080, 40%; chi2 1·6, p=0·29). All 13 CYP who died of SARS-COV-2 with neurological comorbidity had complex neurodisability due to a combination of an underlying genetic or metabolic condition, hypoxic ischaemic events or prematurity. Eight (32%) of the 13 CYP who had a neurological comorbidity also had a respiratory comorbidity, including five who required home respiratory support; four with non-invasive ventilation or high flow oxygen and one with low flow oxygen. There were zero CYP who died of SARS-CoV-2 that were invasively home ventilated. There was one death in a young person with a tracheostomy required for airway patency.

Amongst the 25 CYP who died of SARS-CoV-2 there was one child with each of the following comorbidities; congenital cardiac, oncological, haematological, obesity, endocrinology and complications of prematurity.

There were no deaths in CYP with the following conditions:

1. An isolated respiratory condition e.g. cystic fibrosis or asthma (three of the CYP with complex neurodisability had a historic diagnosis of asthma, however the asthma diagnosis was not considered to contribute to death.)
2. Type 1 diabetes
3. Trisomy 21
4. Isolated diagnosis of epilepsy

The estimated mortality rate for CYP who died of SARS-CoV-2 with a life-limiting condition was 11·5 per 100,000 (95%CI 5·6-21·2) compared to 1,124 per 100,000 (95%CI 1054-1197) for all other causes of death. Although the proportion of CYP with life-limiting neurodisability who died of SARS-CoV-2 was higher, their absolute risk of death was 88·9 per 100,000 (95%CI 47·3-152) compared to 2,441 per 100,000 (95% CI 2,194-2,707) in CYP with life-limiting neurodisability who died of all other causes.

### Place of death

Nine (36%) of the 25 CYP who died of SARS-CoV-2, died within a Paediatric Intensive Care Unit and four died on a hospital ward. The remaining 12 CYP died either at home (unexpected (n=6) or expected (n=2)) or in the Emergency Department (n=4). There were five deaths in CYP with advance care plans in place to provide hospital ward level care rather than escalate to intensive care.

## Discussion

We used a high quality unique national mortality dataset linked to national hospital and SARS-CoV-2 PHE testing data, in-conjunction with clinical review, to identify 25 CYP who died of SARS-CoV-2 infection during the first pandemic year. This corresponds to 2 deaths per million across the CYP population in England. We estimated the infection fatality rate is 5 per 100,000 indicating >99·995% of CYP recover from SARS-CoV-2 infection. SARS-CoV-2 contributed to 0·8% of the 3105 deaths from all causes. During the same time period studied there were 124 deaths from suicide and 268 deaths from trauma, emphasising COVID-19 is rarely fatal in CYP.

This is the first study to differentiate between CYP who have died of SARS-CoV-2 infection rather than died with a positive SARS-CoV-2 test as a coincidental finding. Our result is 60% lower than the figures derived from positive tests thereby markedly reducing the estimated number of CYP who are potentially at risk of death during this pandemic.^22^

The CYP who died of SARS-CoV-2 were more likely to be teenagers than younger children, reflecting the striking continuum of risk increasing through the life-course from infancy to older adult life^23^. Higher proportions of Asian and Black CYP died of SARS-CoV-2 compared to all other causes of death, although deaths were still extremely rare.

Our findings emphasise the importance of underlying comorbidities as the main risk factor for death, as 76% had chronic conditions, 64% had multiple comorbidities, and 60% had life-limiting conditions. The comorbidity group at highest risk were those with complex neurodisability, who comprised 52% of all deaths in CYP who died of SARS-CoV-2. CYP with combined neurodisability and respiratory conditions (8 of the 13 deaths with neurodisability) may be at particularly high risk. CYP with a life-limiting neurodisability have a higher background mortality rate than the general population.^15,24^ There are around 500 deaths annually in this group and therefore SARS-CoV-2 contributed to only 3% during the pandemic. Similarly, for all other comorbidity groups, those who died of SARS-CoV-2 represented a very small proportion of all deaths during the pandemic year. It is important to note we observed no deaths in groups who have been considered at higher risk of respiratory infections, such as CYP with asthma, cystic fibrosis, type 1 diabetes or trisomy 21.

Six CYP who died of SARS-CoV-2 had no evidence of an underlying health condition. This contrasts with other studies which have only reported deaths in CYP who have comorbidity.^6,25^ It is possible, due to the hospital data only being available for the last five years, that some CYP may have had a comorbidity that was not identified in this linkage. It is also possible that CYP in our study had an undiagnosed genetic predisposition to severe disease with SARS-CoV-2 infection.^26^

Our findings extend previous more limited reports on deaths due to SARS-CoV-2 in the UK.^6,7,2525^ The International Severe Acute Respiratory and emerging Infection Consortium (ISARIC) study reported six deaths from 651 admissions across 138 hospitals up to July 2020.^25^ All six CYP had “profound comorbidity” which included neurodisability, extreme prematurity, malignancy and sepsis; three were infants under 28 days of age and three aged 15-18 years.^25^ The methodology in our study enabled demonstration that zero neonates died of SARS-CoV-2 highlighting the value of the clinical review we undertook to determine the role of SARS-CoV-2 in death.

The current UK advice on those defined as “clinically extremely vulnerable” was initially extrapolated from adult risk and it remains very cautious.^2222^ Even taking into consideration the effect of shielding (as both adults and CYP shielded at times during this period) the risk of serious outcomes from SARS-CoV-2 for under 18’s remains extremely low. The risk of removal of CYP from their normal activities across education and social events may prove a greater risk than that of SARS-CoV-2 itself.

The data analysed in this study largely relied upon the quality of the data entered through the NCMD death reporting process. Data completeness was variable, depending on stage of the child death review process. Where possible, we overcame this through discussion with reporting clinicians and data linkage. Rapid data linkage methods were undertaken utilising NHS number alone so this may have resulted in some CYP not being matched to their hospital data. As there is no diagnostic test for PIMS-TS and coding was a challenge it is possible that there may be omissions due to the methods of diagnosis and reporting.

## Conclusion

25 CYP died of SARS-CoV-2 during the first pandemic year in England, equivalent to an infection fatality rate of 5 per 100,000 and a mortality rate of 2 per million. Most had an underlying comorbidity, particularly neurodisability and life-limiting conditions. The CYP who died were mainly >10 years and of Asian and Black ethnicity, compared to other causes of the death, but their absolute risk of death was still extremely low.

These findings are important for guiding policy on vaccination strategies amongst CYP and on guiding families and schools on protection of those at higher clinical risk. Going forward, linkage of the NCMD to other national datasets will enable complete capture of co-morbidities in CYP.

## Data Availability

The data are not available as they contain sensitive identifiable data relating to children who died. Please contact the authors for more information

## Acknowledgements

We would like to thank the three Consultant reviewers (Peter Fleming, Grace Rossouw, and Dorothy Alison Perry) who independently reviewed clinical case notes of the children and young people who died with a positive SARS-CoV-2 test.

We are grateful to the ‘Child Death Overview Panels (CDOP)’ for their support and expertise and all child death review professionals for submitting data and providing additional information when requested. The entire ‘National Child Mortality Database (NCMD)’ team (particularly Nick Cook, Sylvia Stoianova, Vicky Sleap and Tom Williams) have been incredibly helpful in providing data for linkage and supporting analysis. ‘NHS Digital’ and ‘NHS England and NHS Improvement’ Children and Young People data support team have provided data to enable this analysis to occur. We thank Public Health England’s Field Service and National Child and Maternal Health Intelligence Network teams, for their collaboration in establishing the real-time surveillance system on child deaths potentially related to COVID-19 and their ongoing support in the daily linkage with the SARS-CoV-2 test results.

We would like to acknowledge support from the National Institute of Health Research (NIHR) through the National School for Public Health Research Programme and the Applied Research Collaboration North West London.

Parent and public involvement is at the heart of the NCMD programme. We are indebted to Charlotte Bevan (Sands - Stillbirth and Neonatal Death Charity), Therese McAlorum (Child Bereavement UK) and Jenny Ward (Lullaby Trust), who represent bereaved families on the NCMD programme steering group, for their advice and support with setting up the real-time child mortality surveillance system at the beginning of the COVID-19 pandemic.

## References

1 Viner R, Ward J, Hudson L et al. Systematic review of reviews of symptoms and signs of COVID-19 in children and adolescents. 2020. Archive Disease Childhood Epub ahead of print: June 2021. doi:10.1136/archdischild-2020-320972

2 Docherty AB, Harrison EM, Green CA, et al. Features of 20 133 UK patients in hospital with covid-19 using the ISARIC WHO Clinical Characterisation Protocol: prospective observational cohort study. BMJ 2020; 369: m1985 doi: 10.1136/bmj.m1985

3 Bhopal SS, Bagaria J, Olabi B, Bhopal R. CYP remain at low risk of COVID-19 mortality. Lancet Child Adolescent Health 2021; 5(5): e12–e3.

4 Davies P, Evans C, Kanthimathinathan HK, et al. Intensive care admissions of children with paediatric inflammatory multisystem syndrome temporally associated with SARS-CoV-2 (PIMS-TS) in the UK: a multicentre observational study. The Lancet Child & Adolescent Health 2020; 4: 669–77.

5 Whittaker E, Bamford A, Kenny J, et al. Clinical Characteristics of 58 Children With a Pediatric Inflammatory Multisystem Syndrome Temporally Associated With SARS-CoV-2. 2020. JAMA; 324(3): 259–269. doi:10.1001/jama.2020.10369

6 Flood J, Shingleton J, Bennett E et al. Paediatric multisystem inflammatory syndrome temporally associated with SARS-CoV-2 (PIMS-TS): Prospective, national surveillance, United Kingdom and Ireland, 2020. https://doi.org/10.1016/j.lanepe.2021.1000752666-7762

7 Odd D, Stoianova S, Williams T, et al. Archives Disease Childhood 2021. Epub ahead of print. Accessed doi:10.1136/archdischild-2020-320899. Accessed 27th June 2021.

8 Prevalence of SARS-CoV-2 in children and young people in England. Data from Office of National Statistics. Available from https://www.ons.gov.uk/peoplepopulationandcommunity/healthandsocialcare/conditionsanddiseases/bulletins/coronaviruscovid19infectionsurveypilot/11june2021. Accessed 18th June 2021.

9 National Child Mortality Database annual report. Available from https://www.ncmd.info/wp-content/uploads/2020/11/Main-Text-FINAL-WEB.pdf. Accessed 20th May 2021.

10 Ethnicity grouping methodology. Available from https://www.ethnicity-facts-figures.service.gov.uk/style-guide/ethnic-groups. Accessed 21st June 2021.

11 Office for National Statistics. Census geography. An overview of the various geographies used in the production of statistics collected via the UK census. 2019. https://www.ons.gov.uk/methodology/geography/ukgeographies/censusgeography#super-output-area-soa. Accessed 21st June 2021

12 National Child Mortality Database (NCMD) deprivation report. Available from https://www.ncmd.info/2021/05/13/dep-report-2021/. Accessed 21st June 2021

13 Herbert A, Wijlaars L, Zylbersztejn A, Cromwell D, Hardelid, P. Data Resource Profile: Hospital Episode Statistics Admitted Patient Care (HES APC). International Journal of Epidemiology 2017. 46, 4, Pages 1093–1093i

14 Hardelid P, Dattani N, Gilbert R. Estimating the prevalence of chronic conditions in children who die in England, Scotland and Wales: a data linkage cohort study. 2014. BMJ open; 4(8).

15 Fraser LK, Miller M, Hain R, et al. Rising national prevalence of life-limiting conditions in children in England. 2012. Pediatrics; 129(4): e923–e9.

16 Public Health England SARS-CoV-2 testing data information https://www.gov.uk/government/publications/coronavirus-covid-19-testing-data-methodology/covid-19-testing-data-methodology-note. Accessed 10th May 2021.

17 NCMD contributions to modifying the investigation protocol for sudden unexpected deaths in CYP to include post-mortem testing for SARS-CoV-2. Available from https://www.ncmd.info/2020/04/07/jar-covid-19/ Accessed 22nd June 2021

18 Public Health England modelling data for number of children and young people who have had SARS-CoV-2 infection in England. https://coronavirus.data.gov.uk/details/download. Accessed 18th June 2021.

19 ONS data for estimated number of children by age living in England, mid 2019 estimate https://www.ons.gov.uk/peoplepopulationandcommunity/populationandmigration/populationestimates. Accessed 20th May 2021.

20 National Child Mortality Database legal basis for collecting personal and confidential data. https://consult.education.gov.uk/child-protection-safeguarding-and-family-law/working-together-to-safeguard-children-revisions-t/supporting_documents/Working%20Together%20to%20Safeguard%20Children.pdf. Accessed 8th June 2021

21 Control Of Patient Information (COPI) regulations provide a legal basis for linking NCMD data with SUS data https://digital.nhs.uk/coronavirus/coronavirus-covid-19-response-information-governance-hub/control-of-patient-information-copi-notice

22 Royal College of Paediatrics and Child Health. COVID-19 - guidance on clinically extremely vulnerable children and young people. 2021. https://www.rcpch.ac.uk/resources/covid-19-guidance-clinically-extremely-vulnerable-children-young-people (accessed 10th March 2021).

23 Williamson EJ, Walker AJ, Bhaskaran K, et al. Factors associated with COVID-19-related death using OpenSAFELY. Nature 2020; 584(7821): 430–6.

24 ‘Make Every Child Count’ Estimating current and future prevalence of CYP with life-limiting conditions in the United Kingdom https://www.togetherforshortlives.org.uk/resource/make-every-child-count/

25 Swann OV, Holden KA, Turtle L, et al. Clinical characteristics of CYP admitted to hospital with COVID-19 in United Kingdom: prospective multicentre observational cohort study. BMJ 2020;370:m3249 doi: 10.1136/bmj.m3249[published Online First: Epub Date]|.

26 Anastassopoulou, C., Gkizarioti, Z., Patrinos, G.P. et al.. Human genetic factors associated with susceptibility to SARS-CoV-2 infection and COVID-19 disease severity. Hum Genomics 14, 40 (2020). https://doi.org/10.1186/s40246-020-00290-4

27 Children and young people receiving treatment for an Oncological condition in England. Data obtained from SUS data (see reference 12) and discussed with Cancer Programme of Care – Specialised Commissioning, NHS England and NHS Improvement. Data provided 17th June 2021.

28 Children and young people with Congenital Heart Disease in England. Data collected as part of National Institute for cardiovascular outcomes research (NICOR). Data provided by Clinical Reference Group Congenital Heart Disease, NHS England, provided 16th June 2021

29 Children and young people with a diagnosis of Epilepsy in England. Available from https://www.england.nhs.uk/wp-content/uploads/2018/09/E09-S-b-Paediatric-Neurosciences-Neurology.pro_.2013.04.v2.pdf. Accessed 17th June 2021

30 Children and young people with a diagnosis of Asthma in England. Estimates provided by National Asthma and COPD Audit Programme (NACAP), Royal College of Physicians (RCP), England https://www.nacap.org.uk/. Provided 16th June 2021.

