## Supplement for "Deaths in Children and Young People in England following SARS-CoV-2 infection during the first pandemic year: a national study using linked mandatory child death reporting data"

**Supplementary material**

1) Markers of deprivation

From residential address data, the Index of Multiple Deprivation (IMD) 2019 were obtained^[1]^. Deprivation was derived using seven main domains (income, employment, education, health (physical and mental), crime, access to housing and services, and living environment)^[2]^. IMD is derived from the Lower-layer Super Output Area (LSOA) of residence. Each LSOA is a small geographical area with a minimum population of 1000 persons and a mean population of 1500. Each LSOA is placed in one of five categories (1 to 5) containing approximately the same number of people across England, with 1 being the most deprived and 5 being the least deprived^2^.

2) Identification of potential PIMS-TS cases

For all reported deaths from March 2020 to February 2021, the NCMD coding team identified cases where PIMS-TS was considered as a diagnosis using the following process:

a) The NCMD team reviewed all deaths in CYP with sepsis and inflammatory conditions to identify possible cases of PIMS-TS.
b) Using data linkage (see above) this was cross checked with the Paediatric Intensive Care Audit Network (PICANet) and the admitted patient care Secondary Uses Service (SUS) data to ensure all possible cases were identified^[3]^. In November 2020 a new code was introduced for PIMS-TS (UO75). Prior to November 2020, PIMS-TS had been primarily coded within SUS using M303 (Kawasaki disease) and R65 (systemic inflammatory response syndrome) and identifying cases early in the pandemic was problematic. PIMS-TS hospital admissions were defined as those occurring after 1^st^ February 2020 with R65, M303 or U075 as a primary diagnosis only, as we were unable to distinguish between secondary diagnoses related to current or historic admissions with R65 and M303.
c) All possible cases were then reviewed by a Paediatric Infectious Disease and PIMS-TS expert, and where necessary were discussed with the local clinical team.
3) Process to identify if SARS-CoV-2 contributed to death

If it was either clearly apparent within the data provided through death reporting to NCMD that SARS-CoV-2 contributed to death or it was documented 1a on the certificate of cause of death^[4]^, the classification ‘SARS-CoV-2 clearly contributed to death’ was applied. If it was not clearly apparent, each case underwent review by three independent senior specialists who were asked to classify each case according to the categories above. The three reviewers comprised one General Paediatric Consultant, one Neonatology Consultant and one Paediatric Intensive Care Consultant. Each Consultant was blinded to the opinion of the other reviewers.

If two or three Consultant reviewers agreed on the same classification then consensus was achieved. If all three Consultant reviewers disagreed, two further Consultants reviewed the case. The cases classified as ‘Unable to say based on the available information’ were discussed with reporting clinicians and professionals involved in the care. Further specialist opinion was sought from Paediatric Oncologists and a Paediatric Infectious Disease expert due to a high (relative) number of cases (9 and 13 cases respectively) and a need to ensure accuracy of interpretation. The cases of acute COVID-19 illness and PIMS-TS went through the same process of classification. The process that was undertaken to determine the role of SARS-CoV-2 in contributing to death, including the process of classification, is summarised in Figure 1.

4) Small number reporting, information governance and legal basis

*Small number reporting*

A statistical risk assessment for this study determined that whilst identification of individuals may be possible the risk of attribute details being disclosed was low and the public benefit of reporting these small numbers outweighed this risk. Given the sensitive nature of these data and our awareness that clinicians and families may recognise personal experience, we met with a clinician or professional involved in the care of each child or young person who died of SARS-CoV-2 infection. We asked the respective clinician or professional to communicate this work directly with the families.

*Information Governance and Legal Basis*

The NCMD legal basis to collect confidential and personal level data under the Common Law Duty of Confidentiality has been established through the Children Act 2004 Sections M-N, Working Together to Safeguard Children 2018^[5]^. The NCMD legal basis to collect personal data under the General Data Protection Regulation (GDPR) without consent is defined by GDPR Article 6 (e) Public task and 9 (h) Health or social care (with a basis in law).

The PICANet legal basis to process personally identifiable data for the purposes of service evaluation, audit and research was approved by the Patient Information Advisory Group (now the Health Research Authority Confidentiality Advisory Group) in 2002 under Section 60 of the Health and Social Care Act (subsequently Section 251 of the National Health Service Act 2006) (reference: PIAG 4-07(c) 2002). This was amended and approved specifically to collect additional data relating to COVID-19 for confirmed and suspected cases.

Current Control Of Patient Information (COPI) regulations provide a legal basis for linking NCMD data with SUS data without consent^[6]^.

References

1. Office for National Statistics. Census geography. An overview of the various geographies used in the production of statistics collected via the UK census. 2019. Available from <https://www.ons.gov.uk/methodology/geography/ukgeographies/censusgeography#super-output-area-soa>. Accessed 21^st^ June 2021
2. National Child Mortality Database (NCMD) deprivation report. Available from <https://www.ncmd.info/2021/05/13/dep-report-2021/>. Accessed 21^st^ June 2021
3. Herbert A, Wijlaars L, Zylbersztejn A, Cromwell D, Hardelid, P. Data Resource Profile: Hospital Episode Statistics Admitted Patient Care (HES APC). International Journal of Epidemiology 2017. 46, 4, Pages 1093–1093i
4. Guidance for doctors completing medical certificates in England. Available from <https://assets.publishing.service.gov.uk/government/uploads/system/uploads/> attachment_data/file/877302/guidance-for-doctors-completing-medical-certificates-of-cause-of-death-covid-19.pdf Accessed 10^th^ March 2021
5. National Child Mortality Database legal basis for collecting personal and confidential data. <https://consult.education.gov.uk/child-protection-safeguarding-and-family-law/working-together-to-safeguard-children-revisions-t/supporting_documents/Working%20Together%20to%20Safeguard%20Children.pdf>. Accessed 8^th^ June 2021
6. Control Of Patient Information (COPI) regulations provide a legal basis for linking NCMD data with SUS data <https://digital.nhs.uk/coronavirus/coronavirus-covid-19-response-information-governance-hub/control-of-patient-information-copi-notice>
